# Providing breastfeeding support during the COVID-19 pandemic: Concerns of mothers who contacted the Australian Breastfeeding Association

**DOI:** 10.1101/2020.07.18.20152256

**Authors:** Naomi Hull, Renee L Kam, Karleen D Gribble

**Author notes:** **Corresponding author:** Naomi Hull, Australian Breastfeeding Association, Level 3 Suite 2, 150 Albert Road, South Melbourne Victoria, Australia.

## Abstract

Concerns of mothers seeking breastfeeding support during the COVID-19 pandemic, and the experiences of Australian Breastfeeding Association (ABA) volunteers who assisted them, were explored via an online survey. Surveys were completed 16^th^ March to 18^th^ of May 2020 and described the COVID-19 related concerns of 340 individuals. One hundred and thirty six mothers (64%) sought support to protect their infants by continuing breastfeeding, increasing milk supply, or restarting breastfeeding. Mothers were commonly stressed, isolated and needing reassurance. Thirty four (10%) raised concerns about COVID-19 and breastfeeding safety. One hundred and twenty nine (61%) informed volunteers they were unable to access face-to-face health services because of fear or unavailability. Most common breastfeeding concerns were related to insufficient milk or weight gain, painful breasts, relactation, and reducing supplemental milk. Volunteers reported mothers were worried stress had reduced milk supply, that milk supply concerns were exacerbated by the inability to weigh infants, and that seeking medical treatment was being delayed. ABA volunteers stated they felt supported and confident assisting mothers while also expressing distress at mothers’ situation. ABA’s role in emergency response should be recognised and national planning for infant and young child feeding in emergencies, must be urgently developed, funded, and implemented.

## INTRODUCTION

On 11 March 2020, the World Health Organization (WHO) declared the spread of the novel coronavirus SARS-CoV-2, a pandemic (Ghebreyesus, 2020). In Australia, the first confirmed case of COVID-19, the disease caused by the virus, was identified on 25 January 2020 (Department of Health, 2020c). While virus spread was initially slow, by 24 March there were 2051 cases (Department of Health, 2020a). In the third and fourth week of March, Australian governments imposed a variety of public health measures to reduce transmission. These included banning international travel, closure of clubs, gyms, and restaurants, limitation of individual movement outside the home, and gathering restrictions. Schools and universities also shifted to remote learning. At the same time, health services reduced in-person antenatal, postnatal, and child health appointments, and cancelled mothers’ groups. The Australian Breastfeeding Association (ABA) remained operational and continued to support women with breastfeeding.

Formed in 1964, ABA aims to provide women with practical mother-to-mother support to enable them to breastfeed their infants and young children (Australian Breastfeeding Association, 2019). ABA is a registered training organisation and its volunteers undertake competency-based training to receive a Certificate IV in Breastfeeding Education (Counselling or Community). ABA has more than 1000 volunteer breastfeeding counsellors and community educators who deliver antenatal breastfeeding education classes, run mothers’ groups, facilitate social media groups, provide text-based support via the ABA LiveChat service, and provide telephone counselling through the ABA National Breastfeeding Helpline. The National Breastfeeding Helpline received more than 70,000 calls in 2018–19, operates 24 hours a day, 365 days of the year, and receives funding support from the Australian Government (Australian Breastfeeding Association, 2019).

WHO made it clear even before the pandemic was declared, that mothers with COVID-19 should be supported to breastfeed their infants (World Health Organization, 2020c). While early research indicated SARS-CoV-2 infection causes serious disease in 20% of people, significant illness in children was rare (Wei et al., 2020; World Health Organization, 2020d). Experience with other respiratory viruses suggested SAS-CoV-2 transmission via breastmilk was unlikely (World Health Organization, 2020c). The role of breastfeeding in protecting infant health required breastfeeding not be impeded (World Health Organization, 2020c).

WHO published detailed guidance on COVID-19 and neonatal care on 13 March 2020 (World Health Organization, 2020a). This guidance stated that infants born to women who are suspected or confirmed as having COVID-19 should be placed skin-to-skin with their mothers immediately after birth, initiate breastfeeding within an hour of birth, be kept proximate to their mothers day and night, and exclusively breastfeed. These breastfeeding practices should be maintained alongside infection prevention and control practices including respiratory hygiene, hand hygiene, and the cleaning of surfaces (World Health Organization, 2020a). WHO also advised that all mothers should receive support to initiate and establish breastfeeding and to manage breastfeeding difficulties from appropriately trained health professionals and lay breastfeeding counsellors (World Health Organization, 2020a).

From the end of the first week of March, ABA volunteers started reporting mothers were delaying ceasing breastfeeding because of COVID-19 and an increasing number of women wanting relactation assistance. It appeared the pandemic might be changing the feeding practices of mothers and the support ABA volunteers were being asked to provide. ABA national management team members decided to launch an ABA volunteers survey to assess whether ABA was providing volunteers, mothers, and other service users with the support they needed during the COVID-19 pandemic.

## METHODS

Data was collected via an online survey using the SurveyMonkey® platform accessed via a password-protected section of the ABA website between 16 March and 18 May 2020. ABA volunteers were sent an email informing them of the survey, and were also given a message about the survey when they logged into the National Breastfeeding Helpline to take calls. They were asked to complete a survey if people contacting them for assistance as an ABA volunteer raised concerns about COVID-19 or mentioned COVID-19 as a reason for seeking support from ABA.

The survey consisted of nine open-ended questions. Respondents were asked to provide non-identifying information about how and when they were contacted, and the concerns of the person seeking support, noting relactation and increasing milk supply as potential areas of concern.They were also asked whether there was anything they would like to share in relation to their volunteer role and COVID-19 such as whether they felt they had the needed knowledge and skills. Respondents could describe more than one contact in a survey completion and could also complete the survey more than once. No identifying information was collected from respondents.

Survey responses were exported into Excel® and coded using conventional content analysis (Hsieh & Shannon, 2005). The process of coding involved the authors each reading all survey responses and two authors independently undertaking open coding to identify the main ideas being expressed. The three authors then discussed the coding outcomes and finalised a coding framework. The full data set was then re-coded by one author using the finalised framework which was then checked by another author. Where there was ambiguity, the authors determined the appropriate code through discussion. Codes were arranged into themes encompassing the data. Since the data was pre-existing, contains no identifying information, and is of negligible risk, the research was determined to be exempt from the need for ethical review by the Western Sydney University Human Research Ethics Committee.

## RESULTS

The online survey was completed 211 times and described the concerns of 340 individual contacts, including 336 mothers, two fathers, and one grandmother, regarding 341 infants and young children, and one pregnant woman. Respondents stated 207 times they were reporting on contacts made via the National Breastfeeding Helpline, 22 times via LiveChat, and 16 times via other means (eg email counselling, group meetings, social media, breast pump hire). Respondents reported on contacts made between 11 March and 18 May 2020. The most common COVID-19 related issues identified by ABA volunteers and their frequency are summarised in Table 1.

**Table 1.**
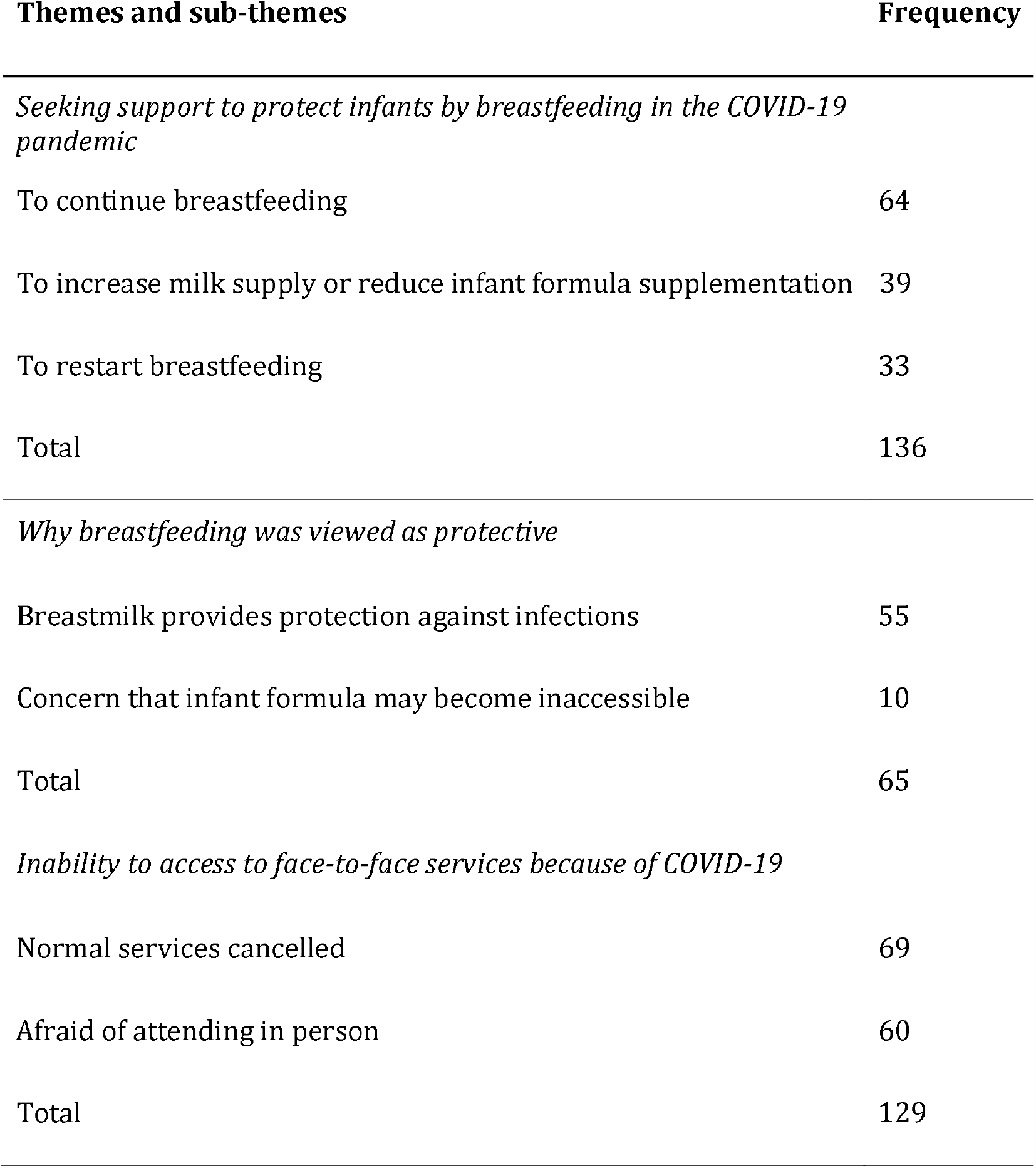
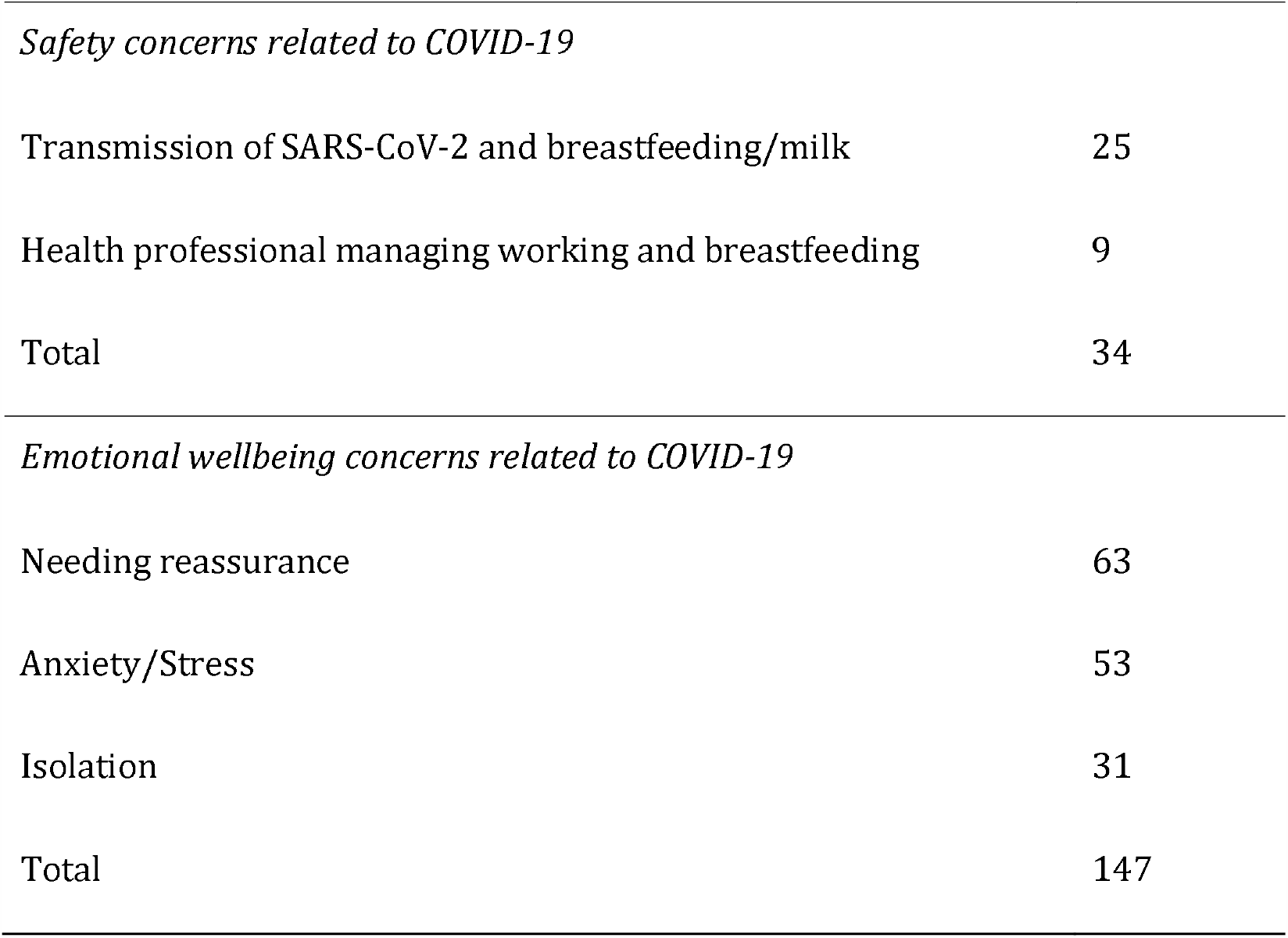
Type and frequency of COVID-19 related issues raised with ABA volunteers.

### Seeking support to protect infants by breastfeeding in the COVID-19 pandemic

One hundred and thirty-six mothers (64% of contacts) were reported to have sought support to continue breastfeeding, to increase their milk supply or to restart breastfeeding to protect their infants.

#### Support to continue breastfeeding

Sixty-four of these women had contacted ABA seeking support to continue breastfeeding or avoid using infant formula. Children of these women ranged from the very young, to older babies and toddlers. For example, ‘*Baby was 12 days old… Had previous poor breastfeeding experiences and happily used formula for other 3 babies. This time mother is concerned about the virus and wants the baby to receive breastmilk’, ‘I had two mothers of newborns under 10 days old. Both had significant anxiety about keeping breastfeeding going to help protect the baby from COVID-19’, and ‘Mother thinking of weaning at 12 months but due to pandemic had decided to continue breastfeeding’*.

#### Support to increase milk supply to avoid or eliminate supplementary formula feeds

Thirty-nine mothers sought assistance to increase their milk supply or reduce infant formula supplementation because of COVID-19. Often infants were young and mothers had been supplementing because of breastfeeding challenges. For example, *‘Nine-day-old baby who had been mostly formula-fed in hospital due to latching difficulties and jaundice wanting to get the baby back onto the breast fully’, and ‘Mother was struggling with attachment, had started… formula but wanted to get breastfeeding right’. There were also instances where reversal of a planned weaning was desired, ‘10-month-old baby. Mother had begun the weaning process, but due to COVID-19, was keen to increase supply so that she could continue to breastfeed’*.

#### Support to restart breastfeeding

Finally, 33 women were reported to have sought assistance with restarting breastfeeding. In some cases, it had been a short time since breastfeeding had ceased and they were reinitiating breastfeeding with a likely existent milk supply, in other cases relactation was necessary. For example, ‘*8 days. Lost weight at birth so formula. Baby willing to breastfeed. Loves it!!’, ‘Mother ceased breastfeeding when babe was 3wks of age. Now 10 weeks…with threat of coronovirus, wants to try to relactate’, and ‘Baby boy 10 weeks old, born premature, mum decided to formula-feed from the start in hospital wanted to know if she could start breastfeeding’*.

### Why breastfeeding was viewed as protective

In almost half of cases (n=65, 48%) where women had sought support to protect their infants by breastfeeding, ABA volunteers recorded why mothers viewed breastfeeding as protective. In the majority of instances (n=55), it was because of the anti-infective properties of breastmilk. For example, ‘Nine day old baby who had been mostly formula fed in hospital…The mother was aware of COVID-19 and wanted the immune system benefits of breastmilk’, *‘Mother wanted to keep her 11 day old breastfeeding to support his immune system for the best protection against the coronavirus’, and ‘Mum has a 9 month old and stopped breastfeeding 6 weeks ago hoping to relactate because she feels breastmilk is important for immune factors with the COVID 19 crisis’*.

A small number of of women (n=10) told volunteers they wanted to breastfeed because they were concerned they might not be able to access infant formula. For example, ‘*Baby 4 weeks old…Mother now wanting to ditch formula and exclusively breastfeed ‘in case he becomes reliant on the formula and I can no longer get it’, and ‘5 1/2-month-old baby highly anxious and teary, worried that if baby continues to refuse and weans anytime over the next few uncertain months that she won’t be able to get formula’*.

### Inability to access to face-to-face services because of COVID-19

An inability to access face-to-face health services was mentioned by 129 (61%) of those who sought support from ABA. In about half of these cases, mothers expressed an unwillingness to go to their General Practitioner (GP) or to see their Maternal and Child Health Nurse (MCHN) because of a fear of exposure to COVID-19. For example, *‘Mother of 3 week old having difficulty seeing her GP and reluctant to see other health professionals, even refused a visit from a maternal and child health nurse because of being worried about COVID-19,’ and ‘Whilst the mother needed help she didn’t want to go out of the house’*.

In the other half of cases, volunteers reported mothers had been unable to access health services because of service reductions. For example, *‘Early discharge due to COVID19 but no associated extra support. This mother was in a rural location with a 1-day-old baby receiving no support at all’, ‘10 day old baby, mother not received any midwife/MCHN visits since being discharged from hospital due to COVID-19’, and ‘Mum with 8-week-old, concerned about her milk supply…she could not get any local assistance from her health service because all their facilities are closed’*.

### Safety concerns related to COVID-19

Volunteers reported 34 instances where contacts raised safety concerns regarding COVID-19. Most concerns were general in nature regarding whether breastfeeding was safe when mothers are infected with SARS-CoV-2. For example, *‘Concerns about feeding her 3 week old baby if she was to contract Covid19 and whether she should wean’*.

In a few instances, the concern was more specific. For example, *‘A mother in NSW who has been tested for the coronavirus and wanted to know if she could continue BF… Very anxious and didn’t want to feed if it would harm her baby’ and ‘Grandmother made contact as her daughter had been advised by care providers at a major tertiary hospital to stop breastfeeding while self-isolating after a possible exposure event’*.

Although relatively small in number (n=9), it was nonetheless striking that health professionals who were breastfeeding themselves, contacted ABA to ask for information. For example, *‘Mum works in healthcare with Covid-19 patients, wanted to know the safety of continuing to express at work v offering baby formula’ and ‘Health care worker* … *preparing to work with COVID-19 cases looking for info about safely cleaning her pumping equipment in a shared sink’*.

### Emotional wellbeing needs

ABA volunteers described 31 mothers who were finding isolation because of COVID-19 difficult, 53 mothers who were anxious or stressed, and 63 instances of mothers needing reassurance or providing mothers with reassurance. For example, *‘Two mothers with babies under 1 week who were feeling very isolated and insecure. I was able to give them information and reassurance’, ‘Was very anxious and stressed as a result of feeling isolated from her mothers’ group and other contacts and felt like she was alone and stuck in ground hog day’, and ‘Feeling very isolated with 10 week old baby. She was very grateful for info re online ABA services’*.

Sometimes volunteers described mothers just wanting to have someone to talk to, *‘Mum feeling isolated and just wanted to talk’, and ‘Many mothers appear to want to talk and be listened to and are on the line for much longer than normal. They just want to talk about the birth, what’s happening with breastfeeding and their baby and their home life’*.

In some circumstances, stress and the need for reassurance was related to being unable to access informal or health professional supports, *‘Mother was very stressed as her normal supports could not help her [gave] reassurance’, and ‘Mother of 6 day old had no in person consults scheduled and had a baby with signs of inadequate intake. Was stressed as did not know how baby would be assessed and cared for appropriately’*.

Some mothers were stressed about the direct threat of COVID-19 and needed reassurance, for example, *‘Both the mothers were very anxious about their babies contracting the coronavirus’, and ‘Both parents COVID positive and in isolation at home. Much reassurance’*.

### Infant feeding concerns raised with ABA volunteers

There were 292 separate breastfeeding concerns raised with ABA volunteers and reported in the survey. The four most common were regarding insufficient milk or inadequate weight gain, painful breasts or nipples, relactation, and infant formula supplementation (Table 2). In many instances, COVID-19 was a factor in why mothers had these concerns and how these concerns presented.

**Table 2.** Most common breastfeeding concerns raised with ABA volunteers.

| Concern | Frequency (n=292*) |
| --- | --- |
| Insufficient milk supply or inadequate weight gain | 80 |
| Painful breast or nipples | 49 |
| Relactation | 27 |
| Reducing infant formula supplementation | 15 |

\*Volunteers could report more than one concern for each individual

#### Insufficient milk supply or inadequate weight gain

Eighty mothers (38%) sought assistance from ABA volunteers because they believed their milk supply or infant’s weight gains were inadequate. Many mothers stated they believed the stress of the COVID-19 pandemic had negatively impacted their milk supply. For example, *‘Mother indicated she felt her supply may have been affected due to her levels of anxiety around COVID’, and ‘Mother was incredibly anxious about the impact of Covid-19. Her partner was a high risk of exposure in his workplace so she was self-isolating with her 4-week-*old … worried that the stress was also affecting her supply’.

Concerns around weight gain were driven or exacerbated by the inability of mothers to access face-to-face health services and to have their infant weighed. For example, *‘Worried about not being able to weigh baby properly as unable to see [health professionals] and using bathroom scales instead’, and ‘Baby 17 days old. Low supply, not sure if baby is getting enough, as baby can’t be weighed. Expressing and supplementing with formula with no extra support’*.

#### Painful breasts and nipples

Forty-nine women (23% of contacts) sought assistance from ABA volunteers with symptoms of mastitis or other breast infections. Many mothers had avoided seeking medical attention for symptoms because of COVID-19. For example, *‘All in relation to mastitis/blocked ducts. They said they would have usually gone to their GP, but they were too scared to because of Covid-19’ and ‘Mother believes she has mastitis (cold chills, breast pain etc over 24 hours) concerned about leaving the house to see GP to get antibiotics’. In one case, access to a doctor was prevented by infection control policies not accounting for mothers having multiple children, ‘Mother had what sounded like a breast infection… I suggested GP visit. She said she had two children and was unable to take both of them to her clinic because they are limited to only 2 people in each consult*.*’ In other instances, concerns regarding the overlap of symptoms between mastitis and COVID-19 jeopardised access to medical care, ‘Mum with mastitis … GP refused to see her in person due to temperature and flu-like symptoms’ and ‘Mum had a blocked duct and was worried that her doctor wouldn’t treat her appropriately if it becomes mastitis, due to the flu-like symptoms… being COVID symptoms’*.

As previously described, concerns related to relactation and reducing infant formula supplementation were connected to mothers wanting to protect their infants in the COVID-19 pandemic.

### ABA volunteer’s feelings about their work

Fifty-nine surveys (28%) included comments from volunteers about their ABA role. As shown in Table 3, these comments most commonly (n=21) reflected that volunteers felt supported and equipped to assist mothers and others who contacted ABA for assistance, *‘I felt I had the knowledge, skills and confidence to answer and reassure them’, and ‘I felt confident my training had stood me in good stead for this crisis’. They sometimes mentioned ABA COVID-19 resources as of assistance, ‘I felt the new resources released by ABA for us helped me to feel more confident answering any inquiries’*.

**Table 3.** ABA volunteers’ feelings about their volunteering work and the resources available to them.

| **Feeling** | **Frequency (n=59)** |
| --- | --- |
| Well supported and equipped to assist | 21 |
| Happy or satisfied | 9 |
| Distressed for mothers | 7 |
| Made suggestions for additional resources | 4 |
| Lacked knowledge | 3 |
| Working beyond capacity | 1 |

Some volunteers (n=9) described being happy and gaining personal satisfaction through helping mothers during the crisis. One reported, *‘It felt good to be able to reassure other mothers in such a challenging time’, and another said, ‘Personally, I felt so happy to be able to do something helpful rather than sitting at home trying not to get infected!’*

In seven surveys, volunteers expressed distress of various kinds at what mothers and infants were experiencing, For example, *‘Feel a bit helpless,’ ‘I am worried about babies falling through the cracks’, and ‘I felt furious that the care providers had said this to the mother’. The feelings of distress were sometimes linked to satisfaction in helping women, ‘I feel really sad for the scared mothers. I feel frustrated that support isn’t getting through to them. I feel so pleased that we exist. I feel able to make a significant difference to mums’. Four volunteers described seeking support for themselves, ‘I am debriefing with my peers and continuing to take Helpline shifts and offer what support I can’*.

Four volunteers suggested resources or actions that could help mothers or themselves including one volunteer who mentioned the need for the National Breastfeeding Helpline to be better advertised, *‘I’d like to see some advertising of our services. The TV mentions Lifeline, Beyond Blue and others but I haven’t heard Helpline at all. Mothers need to know we’re here and waiting for them’. Three volunteers said they lacked knowledge, for example, ‘I wish I’d known more about what GPs are doing to minimise risk and meet the needs of breastfeeding women’. Finally, one volunteer noted she was working beyond her capacity, ‘ATM I am volunteering around 6 hours a day to support our volunteers through training/helpline/online because I know everyone needs extra support right now. This is not sustainable’*.

## DISCUSSION

The findings of this study suggest that the COVID-19 pandemic may have increased the perceived importance of breastfeeding and altered the infant feeding practices of Australian women. Australian Infant Feeding Guidelines recommend infants be fed only breastmilk for the first 6 months and continue breastfeeding into the second year of life (National Health and Medical Research Council, 2012). Ninety-three percent of Australian women initiate breastfeeding but premature cessation of exclusive and any breastfeeding is common (Australian Bureau of Statistics, 2019). In the early postnatal period, as many as 22% of infants are fed supplementary infant formula and a further 16% cease breastfeeding entirely (Ogbo et al., 2017). By 1 year of age, only 40% of infants receive any breastmilk (Australian Bureau of Statistics, 2019). However, our study shows, because of the COVID-19 pandemic, mothers who had not initiated or had stopped breastfeeding, or who were supplementary formula-feeding contacted ABA for assistance to start/restart breastfeeding or increase their milk supply. Infectious disease risk and concerns regarding infant formula availability motivated women to breastfeed. It was particularly notable how many women sought assistance with relactation. Under normal circumstances, support for relactation is an uncommon reason for women to contact ABA. The 2019 ABA client survey (n=395) included only three mothers who nominated relactation as their contact reason. However, the heightened importance of immune protection and food security provided by breastfeeding was a motivation for mothers to restart breastfeeding in the COVID-19 pandemic (Salmon, 2015; Victora et al., 2016).

With the exception of relactation, the most common breastfeeding concerns related to COVID-19 were not different from those encountered under non-emergency conditions. Sore nipples or breasts are consistently the most common concern reported in the annual ABA client survey. Similarly, concerns about low milk supply (which may include concern about weight gain or a desire to decrease formula supplementation) have been in the top four concerns of mothers contacting ABA over the last 3 years. However, lack of access to health care because of COVID-19 had an impact on how these concerns presented. Many mothers described experiencing breast pain and other symptoms likely caused by mastitis but were unable or afraid to visit their doctor to obtain treatment. If left untreated, mastitis can result in tissue destruction and development of abscesses requiring hospitalisation and surgery (Boakes, Woods, Johnson, & Kadoglou, 2018). Anecdotally, La Leche League Italia reported an increase in breast abscesses during the COVID-19 wave of March–April 2020 because mothers had similarly delayed medical treatment (A. Venturini, personal communication, July 10, 2020).

Concerns about milk supply and infant weight gain were also exacerbated by lack of health care access and the inability of infants to be weighed. Similar concerns associated regarding lack of ability to weigh infants has been reported by the Association of Breastfeeding Mothers in the UK (E. Pickett, personal communication, July 17, 2020). Feeding infant formula because of unresolved milk supply concerns would adversely impact infant health (Quigley, Carson, Sacker, & Kelly, 2016). Unidentified dehydration or failure-to-thrive could have serious negative consequences (Jang et al., 2020; Livingstone, Willis, Abdel-Wareth, Thiessen, & Lockitch, 2000; Payne & Quigley, 2016). In two hospitals in Italy, an 84% reduction in paediatric presentations and a 75% decline in admissions occurred in March and April of 2020 raising concerns that fear of seeking medical treatment could imperil child health (Manzoni, Militello, Fiorica, Manzionna, & Cappiello). It seems possible fear or inability to access health care because of COVID-19 could be affecting mothers and infants in many contexts.

Some mothers were concerned stress had affected their milk supply. The erroneous belief that stress reduces milk production is common and, in emergencies, women often perceive milk insufficiency (Gribble, 2014). Changes in infant behaviour such as increased fussiness, more frequent feeds, night waking, and an intolerance of maternal separation are commonly interpreted by mothers as signifying milk supply problems. However, none of these behaviours are reliable indicators of insufficient milk. Rather, such behaviour change is most likely in response to altered circumstances and maternal distress in emergencies and is to be expected. Stress can result in some temporary slowing of oxytocin release and milk flow (but not milk production) also making infants fussy at the breast (Ueda, Yokoyama, Irahara, & Aono, 1994). Breastfeeding counselling can enable women who believe their milk supply is reduced due to stress, to regain confidence in their ability to breastfeed and avoid infant formula use (UNICEF, 2008). For this reason, international infant feeding in emergency guidance requires that emergency plans include provision for breastfeeding counselling (IFE Core Group, 2017).

Inability to access health care, isolation from others, and concerns directly related to COVID-19 were difficult for many mothers. Women were often in need of not only practical assistance and information regarding breastfeeding but reassurance and someone to talk to. The year following the birth of a child is a time of transition as women experience changes to their body, the physical and emotional demands of infant care, and changes in relationships, self-perception, and employment (Fahey & Shenassa, 2013). During this time, social support assists women to develop parenting self-efficacy and facilitates good mental health (Leahy-Warren, McCarthy, & Corcoran, 2012). The pandemic reduced mothers’ access to formal support mediated by the health system and informal support from friends and relatives. Research from Belgium found high rates of symptoms of anxiety and depression in new mothers during COVID-19 lockdown (Ceulemans, Hompes, & Foulon, 2020). Contact with ABA provided women with a form of social support as well as other components of breastfeeding support, like emotional support and reassurance, practical assistance, and informational support (McFadden et al., 2017). The mother-to-mother support ABA provides, may have been particularly salient as volunteers had the shared experience of isolation from family and friends, often were also caring for infants and young children, and all had breastfed themselves (Rossman et al., 2011). Proactive telephone support by ABA counsellors increases breastfeeding rates at 6 months of age (Forster et al., 2019). However, it may be the emotional support provided by volunteers was especially significant to the wellbeing of mothers who were isolated because of COVID-19.

ABA launched the survey reported on in this study partly in order to ascertain whether volunteers were adequately supported. It was reassuring that the majority of those who commented on their volunteer role, stated they felt supported and confident. Their comments noted their general training and the specific ABA COVID-19 resources as of assistance. In March and April 2020, ABA produced COVID-19 resources for volunteers, parents, and health professionals.

Resources regarding the safety of breastfeeding and COVID-19 were targeted at all groups and based on WHO guidance (World Health Organization, 2020a, 2020b). Support materials for volunteers focused on accessing government-funded telehealth services (Department of Health, 2020b), and relactation. A video for health professionals on assessing infant milk intake during telehealth consultations without the ability to weigh was produced (Australian Breastfeeding Association, 2020).

Volunteers reported feeling distressed for mothers and satisfaction in providing assistance. Cultivation of empathy with mothers is core to breastfeeding counselling (World Health Organization, 1993). However, empathic response to individuals in difficulty can lead to helpers experiencing traumatic stress (Regehr, Goldberg, & Hughes, 2002). Alternatively, where assistance is effective, it can lead to compassion satisfaction (Wagaman, Geiger, Shockley, & Segal, 2015). Comments from ABA volunteers reflected some had experienced compassion satisfaction but the distress and workload pressure noted by others underlines the importance of supporting volunteers. It is noteworthy that female ABA volunteers were undertaking their ABA work often alongside paid employment, and a disproportionate burden of household management, childcare, and education supervision noted as impacting women in the pandemic (McLaren, Wong, Nguyen, & Mahamadachchi, 2020).

This research highlights the role of ABA in Australia’s COVID-19 health response. Particularly during the early stages of the pandemic, health service provision suffered as major adjustments had to be made. However, ABA service design meant support to mothers continued largely uninterrupted during the March– May lockdown. ABA volunteers staffing the National Breastfeeding Helpline and LiveChat work from home and no adjustments for isolation measures were necessary. ABA has run online mother support groups (particularly in remote or rural areas) for many years and ABA support groups moved more broadly to online provision from 17 March. Demand for services increased; in April 2020, the National Breastfeeding Helpline received 500–1000 calls in excess of that expected. It is undoubtedly the case that ABA’s services provided a safety net that helped protect mothers and infants. However, as noted by one survey respondent, lack of promotion of ABA’s services may have resulted in under-utilisation. It may have also left a gap for commercial expoitation. The infant formula manufacturer (Nutricia) targeted doctors with advertisements for their helpline stating they were ‘here to help … share the load’ during the crisis. Doctors were invited to direct patients with feeding concerns to their helpline which begins each call with an advertisement for their infant formula store. The exploitation of the pandemic by the baby food industry appears to be an international phenomenon (Cullinan, 2020) which Australia is not immune to.

Despite the role ABA played in supporting mothers and infants during the COVID-19 pandemic, ABA is not mentioned in any Australian emergency plans. More broadly, this pandemic has highlighted again that Australia lacks emergency planning for infants. The National Pandemic Influenza Preparedness Plan recognises infants as a vulnerable group and advises specific strategies be implemented to protect them (Department of Health, 2019). However, neither it nor any state or territory pandemic plan contains infant feeding content. A recent audit of Australian emergency plans and guidance revealed a dearth of planning for infants at all levels of government and in all emergency types (Gribble, Peterson, & Brown, 2019). Nonetheless, the Australian Government’s funding support for the National Breastfeeding Helpline was a measure that contributed to resilience in Australia’s health systems in the COVID-19 pandemic to the benefit of mothers and infants. The Council of Australian Governments’ National Breastfeeding Strategy states that skilled breastfeeding support should be available to mothers and infants during emergencies and that a national policy on infant and young child feeding in emergencies should be developed. The shortcomings of the emergency response in the 2019–20 Black Summer of Bushfires and the COVID-19 pandemic has made it clear such planning must be urgently developed, funded, and implemented.

### Study Limitations

It is a limitation of this study that not all ABA volunteers who were contacted by individuals with concerns related to the COVID-19 pandemic may have completed the study survey. In addition, not all individuals whose breastfeeding concerns were related to the pandemic may have mentioned this to ABA volunteers or fully explained their concerns.

## CONCLUSION

Mothers are responsive to awareness of heightened risks to infants in emergencies and are willing to change feeding practices as a result. Exclusive and continued breastfeeding provides infants with a safe and secure food and water supply and protection from infection and should be promoted as an emergency preparedness activity promoting community resilience. Mothers were negatively impacted by the reduction or removal of formal and informal supports for breastfeeding and mothering during the pandemic. However, ABA services continued to provide support and were able to rapidly deploy key information to mothers, ABA volunteers, and health professionals. The role ABA can play in emergency response should be recognised and reflected in emergency planning. There is an urgent need for the Federal Department of Health to convene and fund a national advisory committee to adapt international infant and young child feeding emergency planning and guidance (IFE Core Group, 2017) for the Australian context.

## Data Availability

Data is not publicly available.

## ACKNOWLEDGEMENTS AND DISCLOSURES

The authors would like to thank Nerida Haines for her work in launching and managing the survey and for supplying ABA statistics for use in the discussion of the paper, the ABA volunteers who completed the study survey, and Libby Salmon for her helpful comments in the development of this paper. No financial assistance or funding was received from any sources towards this article.

